# Incidence rates and symptomatology of community infections with SARS-CoV-2 in children and parents: The CoKids longitudinal household study

**DOI:** 10.1101/2021.12.10.21267600

**Authors:** MLA de Hoog, JGC Sluiter-Post, I Westerhof, E Fourie, VD Heuvelman, TT Boom, SM Euser, P Badoux, C Reusken, LJ Bont, L Sanders, VWV Jaddoe, BL Herpers, D Eggink, JG Wildenbeest, L Duijts, MA van Houten, PCJL Bruijning-Verhagen

## Abstract

**AIM:** The CoKids study aimed to estimate the community incidence of symptomatic and asymptomatic SARS-CoV-2 in children and parents and to assess the symptomatology of SARS-COV-2 infections relative to SARS-CoV-2 negative respiratory episodes.

**METHODS:** In this prospective study, households with at least one child <18 years were recruited from three existing Dutch cohorts. Participation included SARS-CoV-2 screening at 4-6 weeks intervals for all household members during 23 weeks of follow-up and active reporting of new onset respiratory symptoms until July 1^st^ 2021. Follow-up was temporarily intensified following new onset respiratory symptoms in a household member or a SARS-CoV-2 positive screening test and included daily symptom recording, repeated PCR testing (nose-throat, saliva and fecal samples) and SARS-CoV-2 antibody measurement (paired dried blood spots) in all household members. Age-stratified incidence rates for SARS-CoV-2 positive and negative episodes were calculated. Symptomatology and disease burden of respiratory episodes were compared by SARS-CoV-2 status and age.

**RESULTS:** In total 307 households were enrolled including 1209 subjects. We detected 64 SARS-CoV-2 positive and 118 SARS-CoV-2 negative respiratory outbreaks. The highest incidence rate was found in children <12 years for SARS-CoV-2 negative episodes (0.93/ person-year (PY); 95%CI: 0.88-0.96). The SARS-CoV-2 incidence in this age-group was 0.21/PY for confirmed only, and 0.41/PY if probable cases were included. SARS-CoV-2 incidence did not differ by age group (p>0.27). Nasal congestion/runny nose, with or without cough and fatigue were the three most prevalent symptom clusters for both SARS-CoV-2 positive and negative respiratory episodes. Among children, no differences were observed in the symptomatology and severity of SARS-CoV-2 positive versus negative respiratory episodes, whereas among adults, SARS-CoV-2 positive episodes had a higher number and severity of symptoms and with a longer duration p<0.001).

**CONCLUSION:** Using active, longitudinal household follow up, we detected a high incidence rate of SARS-CoV-2 infections in children that was similar to adults. The findings suggest that after 20 months of COVID-19 pandemic, up to 2/3 of Dutch children < 12 years have been infected with SARS-CoV-2. Symptomatology and disease severity of SARS-CoV-2 in children is similar to respiratory illness from other causes. In adults, SARS-COV-2 positive episodes are characterized by more and prolonged symptoms, and higher severity. These findings may assist decisions on COVID-19 policies targeting children.

## INTRODUCTION

The Coronavirus Disease 2019 (COVID-19) pandemic remains a global health crisis with over 265 million confirmed cases reported globally as of December 5^th^ 2021.[1] Contrary to what is observed for many other respiratory virus infections (e.g. RSV, and Influenza), children typically experience less severe disease following a SARS-CoV-2 infection compared to adults. It is estimated that 20-50% of SARS-CoV-2 infections in individuals < 18 years of age occur without symptoms. [2,3] The true incidence of SARS-CoV-2 infections in children may therefore be underestimated. In addition, how the symptomatology and incidence of SARS-CoV-2 infections compares to that of acute respiratory illness (ARI) due to other infections in children has not been systematically quantified. Through intensive, longitudinal monitoring of households with children of different ages, the CoKids study aimed to estimate the community incidence of symptomatic and asymptomatic SARS-CoV-2 infections and to assess the symptomatology of SARS-CoV-2 positive relative to SARS-CoV-2 negative ARI. The study was conducted during the period when children or adolescents were not yet vaccinated against COVID-19. Vaccination of adults was initiated during the final months of the study.

## METHODS

### Study design

Within this prospective cohort study in The Netherlands, households who participated in three existing Dutch birth cohort studies, the RESCEU cohort (current age of children 0-3 years) [4], the MUIS-cohort (current age of children 6-8 years)[5] and the Generation R (current age of children 14-17 years)[6,7], were recruited. Households with one or more children aged 0 to 17 years were eligible to participate in the study. This study was reviewed and ethically approved by the Medical Ethical Committee of the Vrije Universiteit university Medical Centre (VUmc), The Netherlands (reference number A2012.901), the Medical Ethical Committee Utrecht, The Netherlands (reference number 17-069/M) and the Medical Ethical Committee of Erasmus MC, The Netherlands (reference number MEC-2020-0609). Written informed consent was obtained from all participating household members and/or their legal representatives.

The core study consisted of 23-weeks longitudinal follow-up of each household for the occurrence of new onset of respiratory symptoms and/or fever, along repeated SARS-CoV-2 screening at four to six week intervals, irrespective of symptoms. Follow-up was temporarily intensified by means of a household outbreak study that was initiated upon new onset of respiratory symptoms and/or fever in the household irrespective of SARS-CoV-2, or upon introduction of SARS-CoV-2 infection in a household detected through screening or official external testing. The outbreak study included daily symptom diaries and repeated sampling for virological and serological testing for all household members. To limit the burden of study procedures for participants, follow-up was prematurely ended after a household had contributed to two outbreak studies or after a confirmed SARS-CoV-2 positive outbreak. Household enrollment ran from August 2020 through February 2021. The core study was extended to allow for additional follow-up of households with SARS-CoV-2 until July 1^st^ 2021. The extended follow-up no longer included repeated SARS-CoV-2 screening and the outbreak study sampling protocol was only initiated for confirmed SARS-CoV-2 infection in the household.

### Study procedures and data collection

At study enrollment, a baseline questionnaire was completed regarding household characteristics, and medical and demographic characteristics of each household member. During a home visit, research staff provided instructions on self-sampling of a combined nose-throat swab (NTS) and saliva sampling by means of Oracol® sponges. Self-sampling was further supported by instruction videos and leaflets delivered together with the sampling materials.

Households were instructed to actively report new onset of respiratory symptoms or fever in any of the household members throughout follow-up. In addition, SARS-CoV-2 screening at 4-6 weeks interval was performed on self-collected NTS, that were PCR tested within 24 hrs. An outbreak study was launched every time that at least one household member reported new onset of respiratory symptoms or fever, or if a screening test (only in core study) or official external test yielded a SARS-CoV-2 positive result.

At the start of household outbreak, NTS were collected within 48hrs of onset of symptoms of the index case from all household members, irrespective of symptoms. All NTS samples were immediately PCR tested for presence of SARS-CoV-2 virus. Follow-up household sampling schemes were stratified based on presence of SARS-CoV-2 in the initial NTS outbreak samples. The full outbreak sampling protocols for SARS-CoV-2 positive and negative outbreaks are summarized in Table 1. This includes frequent saliva and fecal sampling for SARS-CoV-2 positive outbreaks, as well as additional NTS and saliva for symptomatic subjects throughout the outbreak period. Saliva and fecal samples were temporarily stored in the participant’s home freezer and subsequently transported to the laboratory on dry ice and stored at −80 degrees Celsius (°C) until further analysis. Monitoring of respiratory symptoms was temporarily intensified using daily symptom diaries for each household member, supplemented with daily symptom severity scores and disease questionnaires for those episodes meeting the case definition for Acute Respiratory Illness (ARI, see below for case definition). The following symptoms were included in the diary; nasal congestion/runny nose, cough, headaches, sore throat, fever, shortness of breath, cold shivers, muscle aches, loss of taste or smell, fatigue vomiting and diarrhea. The outbreak study lasted at least 21 days or until 21 days following the last ARI onset in a household member (whichever came later). In addition, all household members collected a dried blood spot by self-finger-prick at the start of the outbreak study and ten days after completion of the outbreak period (convalescent sample) to assess the presence of antibodies against SARS-CoV-2.

**Table 1:**
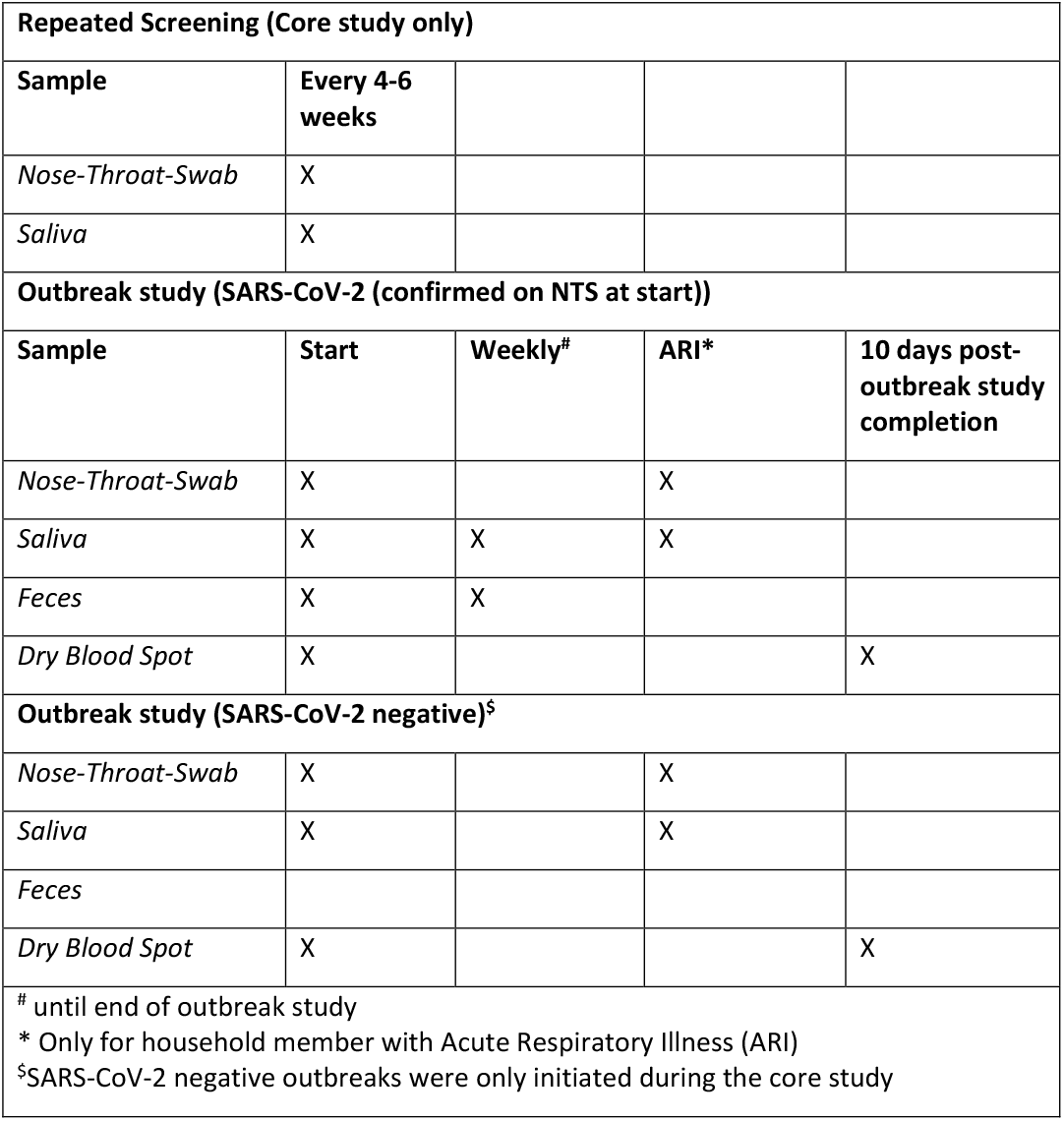
Detailed study sampling scheme for all subjects enrolled.

In the extended follow-up, the outbreak study entry criteria were slightly adapted (table 1); immediate NTS results were only available for the person(s) who reported the respiratory symptoms (index case). If positive, the outbreak study was initiated.

For study procedures and data collection we used a custom made study app (COVapp), compatible with Apple and Android systems, developed by the University Medical Center Utrecht (UMCU) in collaboration with YourResearch Holding BV. The study app was used to collect four types of self-reported data: (1) baseline questionnaire data, (2) reporting of new-onset of respiratory symptoms or fever, (3) registration of all virological and serological specimens collected through participant self-sampling, (4) symptom diaries and disease questionnaires in case of an outbreak. All data entered in the App by the participants were stored in an online secured database and could be accessed and navigated in real-time by study personnel using an online portal with authorized login.

### Laboratory analyses

NTS were analysed using the SARS-CoV-2 RT-PCR based on the presence of the E-gene,[8] which was carried out after a lysis step of the samples. Crossing point (Cp)-values were calculated on Lightcycler 480 1.5.1 software (Roche diagnostics, Basel, Switzerland). This was done by the Regional Public Health Laboratory Kennemerland, Haarlem, The Netherlands.

SARS-CoV-2 presence and viral loads in saliva and fecal samples were analysed by the laboratory of the National Institute for Public Health and the Environment (RIVM), Bilthoven, the Netherlands. The Oracol® sponges were processed according to the manufacturer’s instructions and saliva of both sponges was pooled before further analysis. Total nucleic acid was extracted from 200 μl saliva using MagNApure 96 (MP96) with total nucleic acid kit small volume (Roche). Equine arteritis virus (EAV) was added to the 275 μl lysisbuffer as internal control and yeast tRNA was added as stabilizer. Total nucleic acid was eluted in 50 μl Tris EDTA buffer.

Of the feces specimens a 5% suspension was made in MEM with Hanks’ salts and penicillin and streptomycin, vortex for 15 seconds and 1 minute centrifuged at 16,000 Relative Centrifugal Force. Two-hundred µl supernatant was mixed with 275 µl MP96 lysis buffer including equine arteritis virus (EAV) internal control and yeast tRNA stabilizer.

RT-qPCR was performed on 5 μl total nucleic acid using TaqMan® Fast Virus 1-Step Master Mix (Thermo Fisher) on Roche LC480 II thermal cycler with SARS-like beta coronavirus (Sarbeco) specific E-gene primers and RdRp probe and EAV as described.[8]

NTS, saliva and fecal specimens with a cycle threshold (Cp-/Ct-values) less than or equal to 40 were defined as SARS-CoV-2 positive.

Dry blood spots (DBS) were transported on dry ice to the laboratory, where they were stored at − 80°C. Two bloodspots of 3/16” (containing approximately 6 ul) of serum were punched out of the filter paper and were incubated in 200 μl Blotto-blockingbuffer containing 5% Surfact-Amps (ThermoFisher Scientific Inc., Rockford, USA) overnight at 4°C to release serum from DBS for a test dilution of approximately 1 in 30. 90 μl of the eluted serum was used and subsequently tested for the presence of IgG antibodies reactive with the SARS-CoV-2 spike trimer, S1 and SARS-CoV-2 N antigens in a protein microarray, in duplicate 2-fold serial dilutions starting at 1:20, essentially as described previously.[9] For each antigen, a 4-parameter log-logistic calibration curve was generated to calculate antibody titers (EC50 value). Raw data were processed with the R 4.04 statistical software as described previously.

### Case definitions

Confirmed SARS-CoV-2 infection was defined as 1) a positive RT-PCR result for NTS, saliva or fecal samples, or a documented positive SARS-CoV-2 test result (antigen or PCR) from an official external source (i.e. municipal testing facility); or 2) SARS-CoV-2 negative serology at start of outbreak, and positive at the end of an outbreak period (i.e. seroconversion); or 3) positive serology at start of an outbreak period and respiratory symptoms in the prior 2 weeks that were not previously reported to the study team.

Probable SARS-CoV-2 infection was defined as an episode of respiratory symptoms (at least one day of fever OR two consecutive days with at least 1 respiratory symptom) without confirmation by RT-PCR or serology, but in the presence of one of the household members with a RT-PCR confirmed SARS-CoV-2 infection in NTS.

A SARS-CoV-2 negative episode was defined as presence of respiratory symptoms while all samples were negative for SARS-CoV-2 during the outbreak period, both for the subject and for all household members.

Episodes of respiratory symptoms, either SARS-CoV-2 positive or negative, were further classified as ‘acute respiratory illness’ (ARI) based on daily symptom reports if there were 1) two consecutive days with at least a) one respiratory symptom (cough, sore throat, cold, dyspnea) and one systemic symptom (headache, muscle ache, cold shivers or fatigue) or; b) two respiratory symptoms; or in case of 2) new onset of fever. For ARI episodes we collected additional data on symptom severity using a daily 5-point severity scale, and a disease questionnaire was completed at the end of the episode detailing healthcare visits and medication.

### Statistical methods

The SARS-CoV-2 incidence rate by age category (<12, 12-17 and ≥18 years) was calculated by dividing the number of SARS-CoV-2 infections by the person time of observation. We calculated rates using only confirmed SARS-CoV-2 infections, and rates including both confirmed and probable SARS-CoV-2 infections. Similarly, we calculated the incidence rate of episodes of SARS-CoV-2 negative respiratory illness for different age-groups. The incidence calculations are based on results of the core study period, because during the extended follow-up SARS-CoV-2 negative outbreaks were no longer studied. In addition, no SARS-CoV-2 screenings were performed during the extended follow-up and outbreak entry was only based on a positive NTS test result of the index case, rather than any household member, reducing the overall sensitivity of the protocol to detect SARS-CoV-2 infections. Next, we described symptom frequency and severity comparing SARS-CoV-2 positive and negative respiratory episodes using data collected during the entire study period. Incidence rates and symptomatology were tested for differences by age-group and SARS-CoV-2 status. Statistical analyses were performed with R version 3.6.2 (R Core Team, Vienna, Austria). P-values <0.05 were considered significant.

## RESULTS

### Study population and households

From August 2020 until February 2021, a total of 307 households including 1209 subjects between the age of 0-69 years old were enrolled in the prospective household study (Table 2). In total 403 children < 12 years of age and 179 children 12-17 year old were enrolled. Twenty-six children were 18 years or older and classified as adults. The national SARS-CoV-2 vaccination program for adults started in January 2021, and for those <65 years in April 2021. Therefore, of 627 adults, 205 (32.7%) were fully unvaccinated for the entire duration of the study, 74 (11.8%) received at least one dose during the core study and an additional 287 (45.6%) during the period of extended follow-up. For 58 (9.3%) adults the vaccination status was unknown. During the core study, a total of 179 outbreaks in 149 households were detected of which, 61 were SARS-CoV-2 positive and 118 SARS-CoV-2 negative. An additional three SARS-CoV-2 positive outbreaks were detected during the extended follow-up. Thirty households encountered two or more outbreaks of respiratory symptoms of which 12 households had both a SARS-CoV-2 positive and a SARS-CoV-2 negative outbreak. In total 157 (51%) households did not report any respiratory symptoms or SARS-CoV-2 infection during the entire follow-up. Household size, gender and age distribution were similar in the households with and without SARS-CoV-2 infection. Sample completeness varied substantially by timepoint (Table 3). Over 90% of NTS and saliva samples were completed at the start of the outbreak, but completeness of subsequent saliva and fecal samples declined to 47-66%, depending on the age-group and timepoints. Day one serological sample results were available for 60-96% subjects and convalescent samples for 53-76%.

**Table 2:** Baseline characteristics of the study population

|  | Total | Households with confirmed SARS-CoV-2 | Households without SARS-CoV-2 |
| --- | --- | --- | --- |
| **Total number of household** | 307 | 64 | 243 |
| **Total number of household members** | 1209 | 255 | 954 |
| **Total outbreaks; n (%)** |  |  |  |
| *Core study* | 179 (98.4) | 61 (95.3) | 118 |
| *Extended follow-up* | 3 (1.6) | 3 (4.7) | - |
| **Number of persons in SARS-CoV-2 outbreaks; n (%)** |  |  |  |
| *Core study* | 557 (97.7) | 242 (94.9) | 335 (100) |
| *Extended follow-up* | 13 (2.3) | 13 (5.1) | - |
| **Household size; n (%)** |  |  |  |
| 2 | 7 (2.3) | 2 (3.1) | 5 (2.0) |
| 3 | 82 (26.7) | 14 (21.9) | 68 (28.0) |
| 4 | 155 (50.5) | 33 (51.6) | 122 (50.2) |
| 5 | 51 (16.6) | 12 (18.8) | 39 (16.0) |
| >5 | 12 (3.9) | 3 (4.7) | 9 (3.7) |
| **Gender; n (%)** |  |  |  |
| *Female* | 638 (52.8) | 132 (51.8) | 506 (53.0) |
| *Male* | 571 (47.2) | 123 (48.2) | 448 (47.0) |
| **Age at inclusion; n (%)** |  |  |  |
| <12 | 403 (33.4) | 89 (34.9) | 314 (32.9) |
| 12-17 | 179 (14.8) | 38 (14.9) | 141 (14.8) |
| ≥18 | 627 (51.9) | 128 (50.2) | 499 (52.3) |

**Table 3:** Proportion sample completeness of SARS-CoV-2 positive and negative household outbreaks.

| **Outbreak (Confirmed SARS-CoV-2)** |  |  |  |  |  |  |  |  |  |  |
| --- | --- | --- | --- | --- | --- | --- | --- | --- | --- | --- |
|  |  | NTS; % | Saliva; % |  |  | Feces; % |  |  | Serology; % |  |
| Age (years) | Total in outbreak; n | Start | Start | Week 1 | Week 2 | Start | Week 1 | Week 2 | Start | End |
| <12 | 91 | 96.6 | 94.5 | 50.4 | 51.1 | 49.5 | 49.5 | 47.3 | 83.5 | 52.7 |
| 12-17 | 38 | 97.4 | 97.4 | 57.9 | 65.8 | 60.5 | 52.6 | 47.4 | 92.1 | 76.3 |
| ≥18 | 130 | 97.7 | 93.4 | 54.6 | 54.5 | 55.4 | 48.5 | 49.2 | 96.2 | 66.9 |
| *All ages* | 259 | 96.9 | 94.4 | 53.7 | 55.0 | 54.1 | 49.4 | 48.3 | 91.1 | 63.3 |
| **Outbreak (SARS-CoV-2 negative)\*** |  |  |  |  |  |  |  |  |  |  |
| <12 | 206 | 98.5 | 96.1 | - | - | - | - | - | 60.2 | 36.9 |
| 12-17 | 22 | 100 | 100 | - | - | - | - | - | 95.5 | 68.2 |
| ≥18 | 240 | 98.8 | 96.3 | - | - | - | - | - | 82.5 | 56.3 |
| *All ages* | 468 | 98.7 | 96.4 | - | - | - | - | - | 73.3 | 48.3 |

\*SARS-CoV-2 negative outbreaks were only initiated during the core study

### Incidence of SARS-CoV-2 positive and negative respiratory episodes

Table 4 shows the incidence of SARS-CoV-2 positive and negative respiratory episodes in the different age groups. Within the 61 SARS-CoV-2 household outbreaks, a total of 128 confirmed SARS-CoV-2 infections were detected during 26,780 person weeks of follow-up. An additional 62 subjects developed symptoms during a SARS-CoV-2 outbreak in the household that could not be confirmed by RT-PCR or serology (probable cases). The overall incidence of SARS-CoV-2 was 0.25 per person-year (PY; 95%CI: 0.21-0.29) for confirmed SARS-CoV-2 only and 0.37 per PY (95%CI: 0.33-0.41) for confirmed and probable SARS-CoV-2 cases combined. Overall, no differences in SARS-CoV-2 incidence rates were found between the different age groups (p>0.27)

**Table 4:** Incidence of SARS-CoV-2 positive and negative infections.

| Age (years) | Follow-up time (weeks) | Confirmed SARS-CoV-2 |  | Confirmed + probable SARS-CoV-2 |  | SARS-CoV-2 negative |  |
| --- | --- | --- | --- | --- | --- | --- | --- |
|  |  | Nr. episodes | Incidence rate/person years (95% CI) | Nr. episodes | Incidence rate/person year (95% CI) | Nr. episodes | Incidence rate/person year (95% CI) |
| <12 | 8,944 | 37 | 0.21 (0.16-0.29) | 71 | 0.41 (0.34-0.49) | 160 | 0.93 (0.88-0.96) |
| 12-17 | 3,952 | 17 | 0.22 (0.14-0.34) | 24 | 0.32 (0.22-0.43) | 18 | 0.25 (0.15-0.35) |
| ≥18 | 13,884 | 74 | 0.28 (0.23-0.34) | 95 | 0.36 (0.30-41.7) | 151 | 0.57 (0.50-0.63) |
| All ages | 26,780 | 128 | 0.25 (0.21-0.29) | 190 | 0.37 (0.33-41.3) | 329 | 0.64 (0.60-0.68) |
| Incidence rates are based on the household outbreaks performed during the core study. |  |  |  |  |  |  |  |

Overall, higher incidence rates were found for SARS-CoV-2 negative respiratory episodes compared to SARS-CoV-2 positive episodes (p<0.001). The highest SARS-CoV-2 negative incidence rate was found in children <12 years of age (0.93/PY; 95%CI: 0.88-0.96). Of all respiratory episodes, 13-29% were SARS-CoV-2 infections (confirmed or confirmed + probable, respectively) in children < 12 years of age, 32-51% in children 12-17 years and 26-37% among adults.

### Characteristics of SARS-CoV-2 positive versus negative respiratory episodes

Among the 138 confirmed SARS-CoV-2 infections detected during the entire study period, 24,6% (34/138) of the cases were asymptomatic (Table 5). The proportion of asymptomatic SARS-CoV-2 infections did not differ between age groups (p=0.71). The overall household attack rate for SARS-CoV-2 household outbreaks was 53.3% (47.0-59.5%) considering only confirmed SARS-CoV-2 cases and 79.5% (74.0-84.2%) including both confirmed and probable cases (Table 6).

**Table 5:** Confirmed and probable SARS-CoV-2 cases detected in 64 SARS-CoV-2 household outbreaks.

| Age (years) | Asymptomatic confirmed SARS-CoV-2 |  | Symptomatic confirmed SARS-CoV-2 |  | Probable SARS-CoV-2 |  | SARS-CoV-2 negative* |  |
| --- | --- | --- | --- | --- | --- | --- | --- | --- |
|  | n=34 | % | n=104 | % | N=68 | % | N=52 | % |
| <12 | 14 | 15% | 29 | 32% | 36 | 39% | 12 | 13% |
| 12-17 | 5 | 13% | 12 | 32% | 7 | 18% | 14 | 37% |
| ≥18 | 15 | 12% | 63 | 48% | 25 | 19% | 27 | 21% |
| All ages | 34 | 13% | 104 | 40% | 68 | 26% | 53 | 20% |
| *Subjects who were asymptomatic and with all samples negative for SARS-CoV-2 during the outbreak. |  |  |  |  |  |  |  |  |

**Table 6:** Household attack rate for 64 SARS-CoV-2 outbreaks

| Age (years) | Total number of persons in outbreak | Attack rate confirmed SARS-CoV-2 |  | Attack rate confirmed and probable SARS-CoV-2 |  |
| --- | --- | --- | --- | --- | --- |
|  |  | n | % (95% CI) | n | % (95% CI) |
| <12 | 91 | 43 | 47.3 (36.8-57.9) | 79 | 86.8 (77.7-92.7) |
| 12-17 | 38 | 17 | 44.7 (29.0-61.5) | 24 | 63.2 (46.0-77.7) |
| ≥18 | 130 | 78 | 60.0 (51.0-68.4) | 103 | 79.2 (71.0-85.6) |
| All ages | 259 | 138 | 53.3 (47.0-59.5) | 206 | 79.5 (74.0-84.2) |

Table 7 and Figure 1 and 2 show the symptomatology and disease severity of SARS-CoV-2 positive and negative respiratory episodes stratified for children and adults. Nasal congestion/runny nose, with or without cough and fatigue were the three most prevalent symptom clusters for both SARS-CoV-2 positive and negative respiratory episodes in both adults and children (Figure 1 A-D). In total 74.6% of SARS-CoV-2 positive adults reported fatigue, which frequently persisted for two weeks or more (Figure 2B). Loss of smell/taste was reported by 39.7% of SARS-CoV-2 positive adults versus 3.3% in SARS-CoV-2 negative episodes. In children, this symptom was uncommon in both groups (7.3% versus 1.7%, respectively). Adults also experienced a higher number of symptoms compared to children (figure 1 A and B), a longer duration of symptoms (figure 2 A and B) and higher severity of symptoms (Table 7; p<0.003).

**Table 7:** Disease characteristics of SARS-CoV-2 positive and negative respiratory episodes by age group.

|  | Symptomatic confirmed SARS-CoV-2 |  |  | SARS-CoV-2 negative |  |  |
| --- | --- | --- | --- | --- | --- | --- |
|  | <12 years<br>n=29 | 12-17 years<br>n=12 | ≥18 years<br>n=63 | <12 years<br>n=160 | 12-17 years<br>n=18 | ≥18 years<br>n=151 |
| Days with symptoms (IQR) | 6 (5-12) | 7.5 (4-25) | 13 (7.5-28) | 8 (4-13) | 6 (4.5-11.5) | 5 (32.5-10.5) |
| Number of symptoms (IQR) | 2 (2-4) | 4.5 (2-8) | 6 (4-8) | 2 (2-4) | 4 (3-6) | 3 (2-4) |
| Max severity score (IQR) | 6 (4-9) | 11 (9-20) | 14 (9-19) | 7 (6-12.5) | 10 (6-14) | 7 (6-11) |

**Figure 1A-D:**
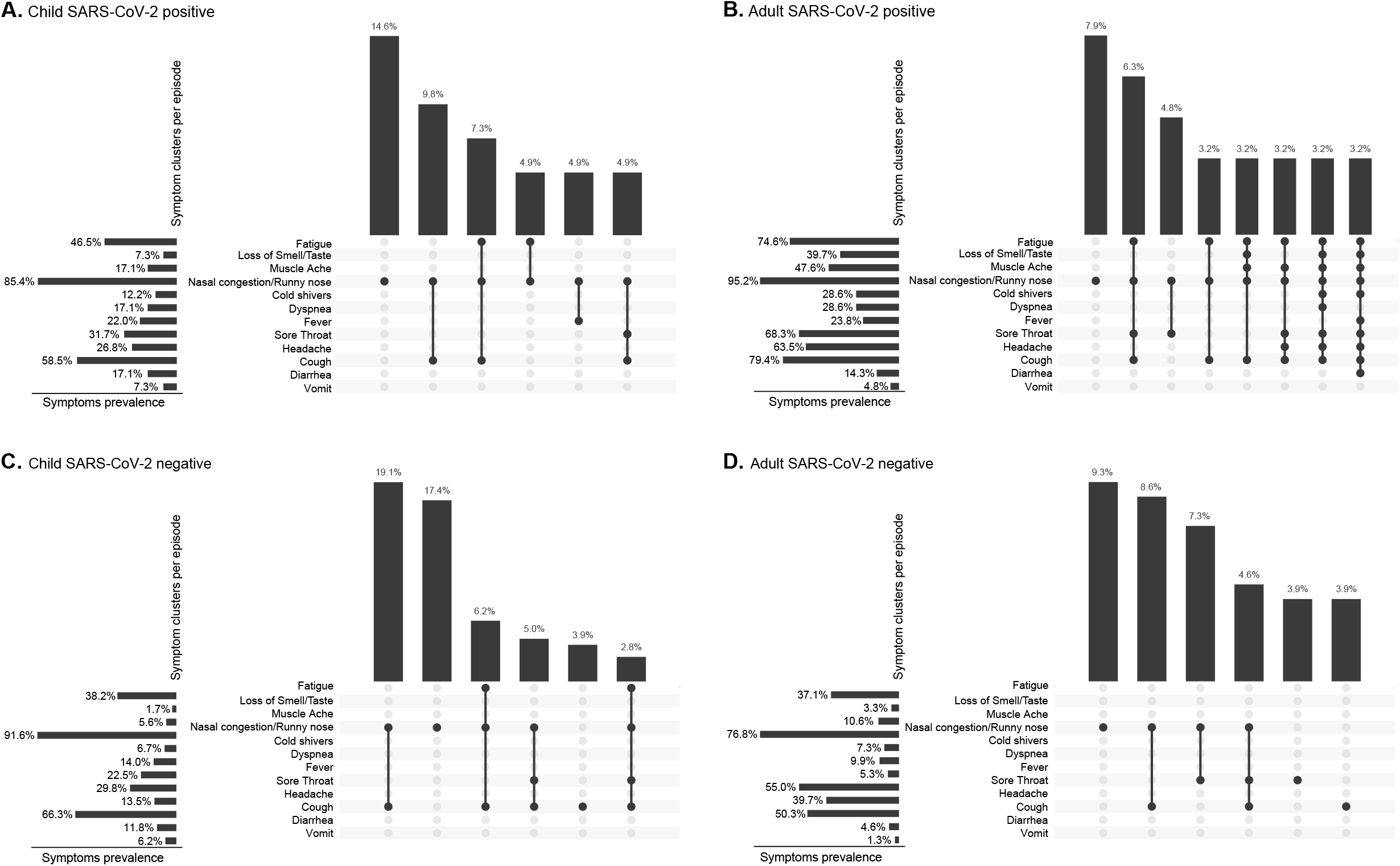
Most common symptom clusters per episode of SARS-CoV-2 positive and negative respiratory episodes in children (<18 years of age) and adults. Footnote: Figure A (n=41), B (n=63), C (n=178) and D (n=151).

**Figure 2A-D:**
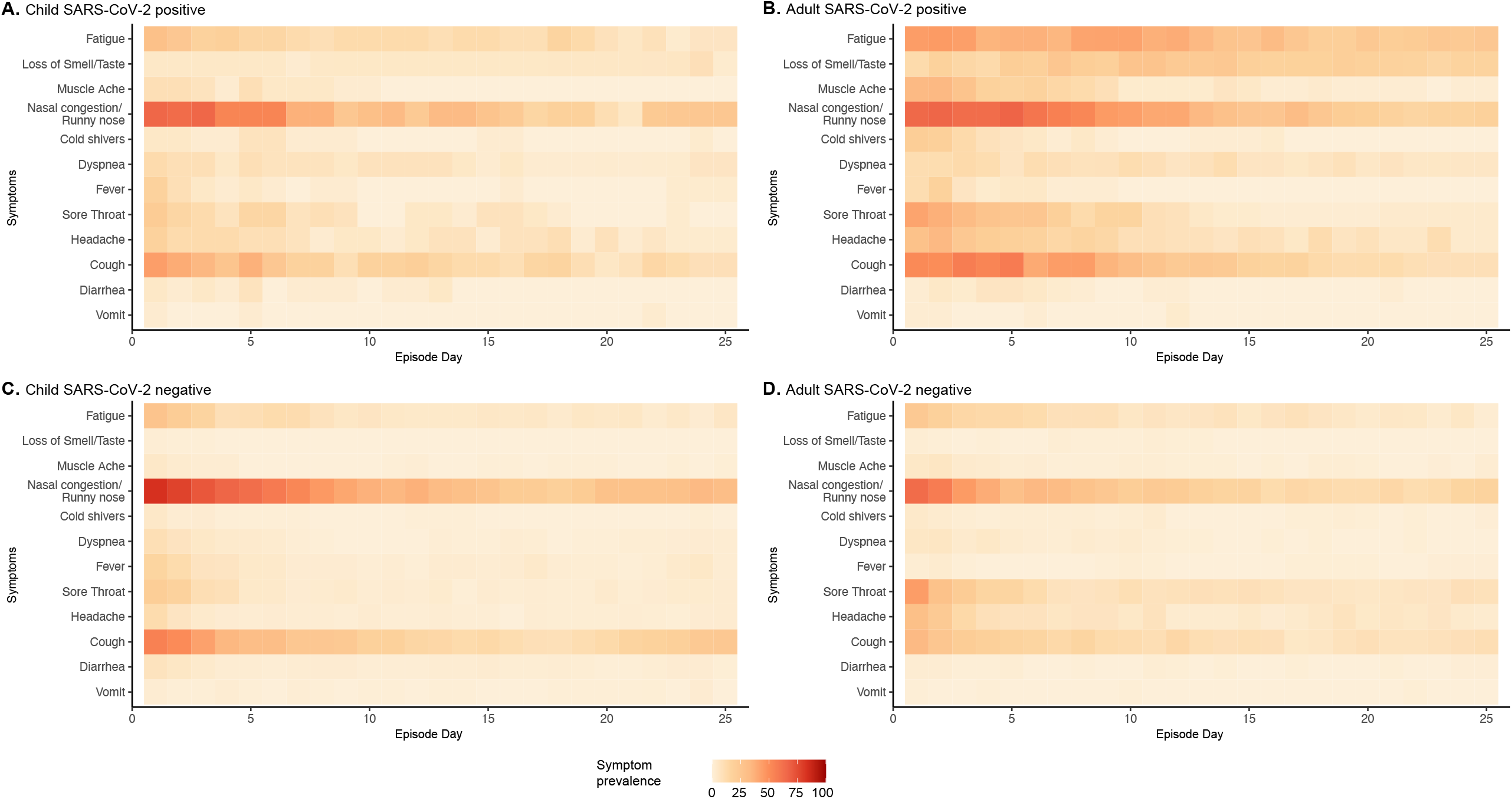
Heat map of individual symptoms up to day 25 of SARS-CoV-2 positive and negative respiratory episodes in children (<18 years of age) and adults.

In children <18 years of age, no clear distinction could be made between SARS-CoV-2 positive versus negative respiratory episodes based on the reported symptoms (Figure 1A and 1C). In adults, generally more respiratory symptoms were reported for a longer duration during a positive SARS-CoV-2 episode compared to a negative SARS-CoV-2 respiratory episode (Figure 1B and 1D and Figure 2A and 2D).

## DISCUSSION

In our study, conducted when adults and children in the Netherlands were largely unvaccinated, we observed that SARS-CoV-2 infection rates did not significantly differ between children and adults, contrary to what has been reported in early studies and in national SARS-CoV-2 surveillance data.[10,11] Younger age-groups generally experienced milder disease or asymptomatic SARS-CoV-2 infection compared to adults and symptoms resemble those of other common respiratory infections in children. Without active, longitudinal follow-up and extensive sampling, many infections in children may therefore go unnoticed. At the incidence rate found in our study of 0.21-0.41 SARS-CoV-2 infections per year, we estimate that 20 months into the pandemic at least one-third and probably up to two-third of children < 12 years have already experienced a first episode of SARS-CoV-2 infection in the Netherlands. These estimates are higher than those obtained from seroprevalence data[12] and therefore complementary to inform vaccine policy decisions, as the direct health benefits from vaccination are expected to be larger for SARS-CoV-2 naïve children than for those previously infected. Furthermore, in children with previous exposure to SARS-CoV-2, reduced dosing schedules could be considered. In the Netherlands, vaccination of children 5-11 years old has not yet started. Our results may help assess the potential impact of vaccination on the pediatric SARS-CoV-2 disease burden in relation to the timing of implementation as infections keep accumulating in this age-group and the proportion of naïve subjects will further decline.

Our study also provides a granular and head-to-head comparison of the symptomatology of SARS-CoV-2 positive and negative respiratory episodes in the community setting. We observed that the symptomatology and severity of disease is very similar for SARS-CoV-2 positive versus negative respiratory episodes in children, whereas among adults, SARS-CoV-2 infections clearly resulted in more symptom burden and prolonged disease compared to SARS-CoV-2 negative episodes. Among children, the most common symptom clusters where runny nose/nasal congestion with or without cough and fatigue for both SARS-CoV-2 positive and negative episodes. This confirms that in most children, it is not possible to distinguish SARS-CoV-2 from other respiratory infections based on symptoms. In adults, loss of smell/taste is a discriminative symptom for SARS-CoV-2, present in 39.7% of SARS-CoV-2 infected subjects versus 3.3% of those testing negative, but in children this symptom was uncommon in both groups (7.3% in positive, versus 1.7% in negative children). Our results also further confirm that disease burden from SARS-CoV-2 in children is generally mild and comparable to that of other respiratory infections. Of note, Influenza Virus and Respiratory Syncytial Virus are known to be associated with more severe respiratory disease in children, but were not circulating during the period when this study was conducted, presumably as a result of the COVID-19 non-pharmaceutical control interventions. This may have lowered the average disease burden of SARS-CoV-2 negative respiratory episodes in children in our study compared to normal seasons.

Our study has some limitations that need to be addressed;

First, despite the extensive sampling protocol, it is possible that SARS-CoV-2 infections were underdetected to some extent due to missing outbreak samples or asymptomatic infections in-between screenings. Sample completeness was high for the initial outbreak samples, but declined thereafter and convalescent serology was only available for 48.8% and 63.3% of subjects in SARS-CoV-2 positive and negative outbreaks, respectively. We therefore believe that the incidence and attack rates reported in this study should be interpreted as minimum values, rather than averages. Similarly, the true proportion of asymptomatic infections could be somewhat higher than estimated in this study.

Second, a large fraction of the observation time fell in the period when the alpha variant of SARS-CoV-2 was dominant in the Netherlands. The period of extended follow-up mostly covered a period with dominance of the Delta variant. Although symptomatology appears not to be different for alpha versus delta, it is possible that household transmission rates were higher during delta dominance, which is only minimally captured in our data (three SARS-CoV-2 outbreaks were studied during extended follow-up). Alternatively, more parents received COVID-19 vaccination during the extended follow-up period and this most likely lowered infection rates among these households. It is unlikely that vaccination had significant impact on the results of the core study, used to calculate incidence rates, as only 74 out of 627 parents received a first or second dose during this period.

Third, this study was conducted in the Netherlands. Incidence rates during the same period may have been different in other countries and settings, depending on the level of epidemic control and measures such as school closures. In the Netherlands, all schools closed mid-December 2020 as part of a full lockdown. Elementary schools reopened early February 2021 and secondary schools opened early March 2021 at reduced student occupancy. In addition, it is possible that the period of full lockdown in Winter 2020/2021 temporarily increased within household exposure and consequently the household attack rate, but this effect has not been confirmed. In summary, the SARS-CoV-2 incidence estimates should be interpreted in the context of the prevailing COVID-19 situation at the time and location of the study.

## CONCLUSION

Using active, longitudinal household follow up, we detected a high incidence of SARS-CoV-2 infections in children of all ages, that is comparable to (unvaccinated) adults, including about 25% asymptomatic infections. These findings suggest that, pending the implementation of COVID-19 vaccination in < 12 years, up to 2/3 of Dutch children have been previously infected with SARS-CoV-2. Symptomatology and disease severity of SARS-CoV-2 in children is similar to respiratory illness from other causes. In adults, SARS-COV-2 positive episodes are characterized by more and prolonged symptoms, and higher severity compared to those testing negative for SARS-CoV-2. These findings may assist decisions on COVID-19 policies targeting children.

## Data Availability

All data produced in the present study are available upon reasonable request to the authors

## AKNOWLEDGEMENT

We gratefully acknowledge the contribution of children and their parents in the study.

We thank Mara van Roermund, Inki Somsen-de Boer, Valentina Pavlova, Michael Arvin, Fenna Geugjes, Eline Westra en Inge van Riessen for their contribution in the data collection of the MUIS and RESCEU Study. We thank Elise Abal van der Pieterman and Lizette van Veen-Berkx for their contribution in the data collection of the Generation R CoKids Study.

We thank Lisa Wijsman; Bas van der Veer; Annemarie van den Brandt; Jeroen Cremer; Sharon van den Brink; Ryanne Jaarsma; Kim Freriks; Lynn Aarts; Sanne Bos, Euníce Then, Sophie van Tol, Gert-Jan Godeke, Alexia Bottino, Hanae Abba, David Gideonse, Fion Brouwer and Johan Reimerink of the National Institute for Public Health and the Environment (RIVM) and the lab researchers of the Regional Public Health Laboratory Kennemerland, for laboratory analyses.

The Generation R Study is conducted by the Erasmus MC in close collaboration with the School of Law and the Faculty of Social Sciences at the Erasmus University, Rotterdam; the Municipal Health Service, Rotterdam area; and the Stichting Trombosedienst and Artsenlaboratorium Rijnmond (Star-MDC), Rotterdam. We gratefully acknowledge the contribution of the general practitioners, hospitals, midwives, and pharmacies in Rotterdam.

The REspiratory Syncytial virus Consortium in EUrope (RESCEU) is conducted by the University Medical Center Utrecht in close collaboration with Spaarne Gasthuis (Hoofddorp, the Netherlands).

The MUIS study is conducted by the Spaarne Gasthuis in close collaboration with the Department of Obstetrics and Gynecology of the Spaarne Gasthuis (Hoofddorp, the Netherlands) and participating midwifery clinics.

The CoKids study is part of the COVID program funded by ZonMw (grant number: 10150062010006) and the RIVM.

## Notes

### Competing Interest Statement

The authors have declared no competing interest.

### Funding Statement

The CoKids study is part of the COVID program funded by ZonMw (grant number: 10150062010006) and the National Institute for Public Health and the Environment (RIVM).

### Author Declarations

The Medical Ethical Committee of the Vrije Universiteit university Medical Centre (VUmc) (RESCUE cohort), Medical Ethical Committee Utrecht, The Netherlands (RESCEU cohort) and Medical Ethical Committee of Erasmus MC, The Netherlands (GenerationR cohort) gave ethical approval for this work

## REFERENCES

[1] COVID-19 Weekly Epidemiological Update: World Health Organisation. Weekly epidemiological update on COVID-19 - 7 December 2021 (who.int). 7 December 2021.

[2] Dawood FS, Porucznik CA, Veguilla V, Stanford JB, Duque J, Rolfes MA, et al. Incidence Rates, Household Infection Risk, and Clinical Characteristics of SARS-CoV-2 Infection among Children and Adults in Utah and New York City, New York. JAMA Pediatrics 2021. https://doi.org/10.1001/jamapediatrics.2021.4217.

[3] Han MS, Choi EH, Chang SH, Jin B lo, Lee EJ, Kim BN, et al. Clinical Characteristics and Viral RNA Detection in Children with Coronavirus Disease 2019 in the Republic of Korea. JAMA Pediatrics 2021;175:73–80. https://doi.org/10.1001/jamapediatrics.2020.3988.

[4] Wildenbeest JG, Zuurbier RP, Korsten K, van Houten MA, Billard MN, Derksen-Lazet N, et al. Respiratory syncytial virus consortium in Europe (RESCEU) birth cohort study: Defining the burden of infant respiratory syncytial virus disease in Europe. Journal of Infectious Diseases 2020;222:S606–12. https://doi.org/10.1093/INFDIS/JIAA310.

[5] Bosch AATM, de Steenhuijsen Piters WAA, van Houten MA, Chu MLJN, Biesbroek G, Kool J, et al. Maturation of the infant respiratory microbiota, environmental drivers, and health consequences. American Journal of Respiratory and Critical Care Medicine 2017;196:1582–90. https://doi.org/10.1164/rccm.201703-0554OC.

[6] Kooijman MN, Kruithof CJ, van Duijn CM, Duijts L, Franco OH, van IJzendoorn MH, et al. The Generation R Study: design and cohort update 2017. European Journal of Epidemiology 2016;31:1243–64. https://doi.org/10.1007/s10654-016-0224-9.

[7] Kruithof CJ, Kooijman MN, van Duijn CM, Franco OH, de Jongste JC, Klaver CCW, et al. The Generation R Study: Biobank update 2015. European Journal of Epidemiology 2014;29:911–27. https://doi.org/10.1007/s10654-014-9980-6.

[8] Corman VM, Landt O, Kaiser M, Molenkamp R, Meijer A, Chu DKW, et al. Detection of 2019 novel coronavirus (2019-nCoV) by real-time RT-PCR. Eurosurveillance 2020;25. https://doi.org/10.2807/1560-7917.ES.2020.25.3.2000045.

[9] van Tol S, Mögling R, Li W, Godeke GJ, Swart A, Bergmans B, et al. Accurate serology for SARS-CoV-2 and common human coronaviruses using a multiplex approach. Emerging Microbes and Infections 2020;9:1965–73. https://doi.org/10.1080/22221751.2020.1813636.

[10] Madewell ZJ, Yang Y, Longini IM, Halloran ME, Dean NE. Household Transmission of SARS-CoV-2: A Systematic Review and Meta-analysis. JAMA Network Open 2020;3:e2031756. https://doi.org/10.1001/jamanetworkopen.2020.31756.

[11] Coronadashboard_RIVM: Rijksoverheid. https://coronadashboard.rijksoverheid.nl/landelijk/positief-geteste-mensen. 2021.

[12] Pienter Corona onderzoek: Rijksinstituut voor Volksgezondheid en Milieu. https://www.rivm.nl/pienter-corona-onderzoek/resultaten. 23 November 2021

